# Non-inferiority of essential medicines for caries arrest and prevention in a school-based program: Results from the Caried*Away* pragmatic clinical trial

**DOI:** 10.1101/2022.04.26.22274321

**Authors:** Ryan Richard Ruff, Tamarinda Barry-Godin, Richard Niederman

## Abstract

**Background:** Dental caries is the most common global childhood disease. To control caries, the Centers for Disease Control and Prevention recommends school-based caries prevention, and the World Health Organization lists glass ionomer cement and silver diamine fluoride as essential dental medicines. The Caried*Away* trial tested the comparative effectiveness of these essential medicines when used in a school-based dental care program.

**Methods:** This cluster-randomized non-inferiority pragmatic trial was conducted in children from 2018 to 2022. Subjects were randomized at the school level to receive either silver diamine fluoride (”simple care”) or an active comparator of glass ionomer sealants and atraumatic restorations (”complex care”). All subjects received tooth brushes, fluoride toothpaste, and fluoride varnish. We assessed caries arrest and incidence at two years using mixed-effects multilevel models and two-sample proportion tests with clustering adjustment.

**Results:** 1398 subjects received treatment and completed follow-up observations after two years. The proportion of subjects with arrested caries in simple and complex groups was 0.56 and 0.46, respectively (difference = -0.11, 95% CI = -0.22, 0.01). Prevention rates for no new caries were 0.81 and 0.82 (difference = 0.01, 95% CI = -0.04, 0.06).

**Conclusions:** Over a two-year, non-intervention period, simple care was non-inferior to complex care for both caries arrest and prevention. Results support the utilization of silver diamine fluoride as an arresting and preventive agent in school-based oral health programs and questions the periodicity of current caries prevention recommendations.

## 1 Introduction

Dental caries (tooth decay) is a natural process by which bacteria in the biofilm cause fluctuations in pH, leading to enamel erosion and a resulting visible lesion [1]. If left untreated, caries can result in pain, abscess, and systemic infection, leading to functional and/or psychosocial impairment [2]. Caries is the most prevalent childhood disease in the world and is most prominent among lowincome populations [3]. The disproportionate burden of caries in vulnerable groups largely stems from a lower dental service utilization; those most at risk often lack access to preventive services or affordable care [4, 5].

To reduce children’s caries burden, multiple state and federal organizations, including the Centers for Disease Control and Prevention, recommend dental sealants and topical fluorides as part of a school-based caries prevention program. Similarly, the World Health Organization lists silver diamine fluoride (SDF) and glass ionomer cement as essential medicines for dental caries [6]. The efficacy of these treatments is well established: topical fluorides prevent caries in primary and permanent teeth when applied at least twice per year [7, 8]; dental sealants significantly reduce caries incidence in primary and secondary molars and arrest the progression of noncavitated lesions [9]; atraumatic restorative treatment (ART) non-invasively arrests caries [10, 11, 12]; and SDF reduces the risk of root carious lesions and controls caries progression [13, 14].

The effectiveness of these preventive options when used in a school-based program is unknown. Here we report on the comparative effectiveness of silver diamine fluoride and glass ionomer in arresting and preventing caries using a school-based, pragmatic, non-inferiority trial design.

## 2 Methods

This study received ethical approval from the New York University School of Medicine Institutional Review Board (i17-00578) and is reported following the CONSORT guidelines for randomized trials. Detailed study information is previously published in an available trial protocol [15]. The study is registered at www.clinicaltrials.gov (#NCT03442309).

### 2.1 Design and Participants

Caried*Away* was a longitudinal, cluster-randomized, single-blind, pragmatic non-inferiority controlled trial to evaluate the effectiveness of silver diamine fluoride with fluoride varnish in comparison to an established, active comparator of glass ionomer sealants/fluoride varnish and atraumatic restorative treatment for the arrest and prevention of dental caries. The study employed a two-stage enrollment process. First, eligible schools in the New York City area were solicited for participation. Inclusion criteria for school enrollment included: an overall student population of 80% or higher receiving free or reduced lunch (as a proxy for low socioeconomic status), at least 50% of enrolled students reporting as Hispanic/Latino or black ethnicities, and the absence of a preexisting dental healthcare provider operating within the school. No restrictions were made on the racial/ethnic distribution of the remaining school population. Second, informational letters and informed consent documents were distributed to all children enrolled in participating schools. Any subject with completed parental informed consent and assent was enrolled in the study. Exclusion criteria for individual subjects included any child who did not speak English and children enrolled in special education classrooms.

### 2.2 Randomization

Consenting schools were listed in ascending order of population size and block randomized in blocks of four schools using a 1:1 allocation ratio. Allocation sequences were created using a random number generator.

### 2.3 Interventions

Subjects were randomized at the school-level to receive either “simple” (experimental group) or “complex” (a standard of care active comparator) treatment. Simple treatment consisted of fluoride varnish (5% NaF, Colgate PreviDent) applied to all teeth and 38% silver diamine fluoride (Elevate Oral Care Advantage Arrest 38%, 2.24 F-ion mg/dose) applied to all asymptomatic cavitated lesions and brushed on all pits and fissures of bicuspids and molars for 30 seconds. Complex treatment included identical application of fluoride varnish, glass ionomer sealants (GC Fuji IX) applied to all pits and fissures of bicuspids and molars, and placement of atraumatic restorations on all frank asymptomatic cavitated lesions. All subjects received tooth brushes and fluoride toothpaste.

Clinical care was provided in a dedicated room in each school using mobile equipment by either dental hygienists or registered nurses with the support of dental assistants and under the supervision of a licensed dentist.

### 2.4 Data Collection

At each observation, study clinicians performed full-mouth visual-tactile oral examinations. Teeth were assessed as being present or missing intraorally. Individual tooth surfaces were assessed as being intact/sound, sealed, restored, decayed, or arrested. All examiners were standardized using identical diagnostic and treatment protocols. Data were recorded on password-protected tablet computers using proprietary electronic health record software.

### 2.5 Caries and Surface Diagnoses

Caries diagnosis was performed using the International Caries Detection and Assessment System’s (ICDAS) adapted criteria for epidemiology and clinical research settings. Individual tooth surfaces were assessed as being intact/sound (ICDAS II codes 0–4), sealed, restored, decayed (ICDAS II code 5–6), or arrested [caries]. Full details of clinical protocols are included in supplementary appendices.

### 2.6 Outcomes

Our primary study outcomes were the proportion of subjects with arrested carious lesions (arrest) and the proportion of subjects with no observed incidence of decayed teeth from previously sound dentition (prevention). Outcomes were calculated at the individual level to avoid within-subject correlation for participants having multiple lesions at baseline or multiple new caries at follow-up. For example, if a subject at baseline presented with multiple carious lesions that received treatment, a failure of any treated lesion at follow-up was considered arrest failure regardless of the arrested status of other lesions. Similarly, any new incidence of decay was considered prevention failure, regardless of how many lesions were observed.

Decay prevalence was defined as any untreated decay on any whole tooth. For caries arrest, teeth with carious lesions at baseline which were treated using either simple or complex care were identified. Each identified tooth was then checked at follow-up. If the tooth presented with decay at follow-up or presented with a filling that was applied by an external clinician, and was not exfoliated and replaced by a permanent tooth (if the tooth was originally deciduous) between visits, it was determined to be arrest failure. The tooth was otherwise coded to be arrested. If a tooth was exfoliated prior to the 24-month follow-up that was coded as arrested at baseline, that tooth was discounted from analysis. For caries incidence, the number of decayed teeth not previously carious or arrested and the number of teeth with new fillings were counted at each visit and a summary score was calculated for each subject at each visit.

### 2.7 Demographic Variables

Demographic information including age, sex, and race/ethnicity were obtained from informed consent documents or school records. A unique identification number maintained by the Office of School Health at the New York City Department Health and Mental Hygiene and New York City Department of Education was similarly used as the patient record number for this study.

### 2.8 Blinding

Participants were blinded to their group assignments, however given the staining effect of SDF on untreated decay, it was possible that patients could derive their treatment assignment. Clinicians and examiners were not blinded due to the specific procedures required for each treatment.

### 2.9 Statistical Analysis

Subjects were analyzed according to intent to treat. Any subjects who switched schools that were randomized to different treatment arms were analyzed according to their original treatment assignment. Power analyses for primary clinical outcomes were previously reported [15]. The intraclass correlation for dependence within cluster was estimated via mixed effects multilevel logistic models. For caries arrest and prevention, our null hypothesis was *H*_0_ : *C − S >*= 10, while our alternative hypothesis was *H*_*a*_ : *C* − *S* < 10. Our statistical test for this hypothesis used two-sample proportion tests adjusting for any clustering effect of schools and by comparing the upper bound of the two-sided 95% confidence interval for (C-S) to the non-inferiority margin [16, 17].

Analysis of arrest included mixed dentition. Each tooth treated with either simple or complex prevention was considered as a single trial with a dichotomous outcome of either of arrested or failed to arrest over the course of observation. Arrest failure was recorded if the tooth presented at baseline with untreated caries, received treatment at baseline with either simple or complex care, and presented at follow-up with either untreated caries or a filling. Any failure of an individual tooth was considered failure at the subject level, regardless if other lesions in the same subject remained arrested. Analysis of caries prevention considered the proportion of subjects in each treatment group that developed new caries post-treatment. New caries for prevention analyses included subjects that presented at follow-up with either (1) untreated carious lesions or (2) presence of a filling that was not present at baseline, indicating development of new disease prior to follow-up.

Analysis was conducted in Stata v17 and R v1.4. Statistical significance was set at 0.05.

## 3 Results

The participant flow diagram for the Caried*Away* trial is presented in Figure 1. Our analytic sample consisted of all subjects who were enrolled, randomized, and treated with a single application of either SDF or sealants/ART and completed a second follow-up visit after approximately two years. A total of 4718 subjects across 47 schools were treated at baseline between September 2019 and March 2020. After removing any subject in any treatment group that aged out of the study, exfoliated teeth treated at baseline, or was otherwise unavailable yielded a sample of 2998. We completed follow-up observations between September 2021 and 2 March 2022 with 1398 subjects (611 in the experimental arm, 787 in the active control), for an overall follow-up rate of 30%. As caries arrest can only be evaluated in subjects that had untreated decay at baseline, the analytic sample for arrest was 413 patients. The analytic sample for prevention was 985 patients.

**Figure 1:**
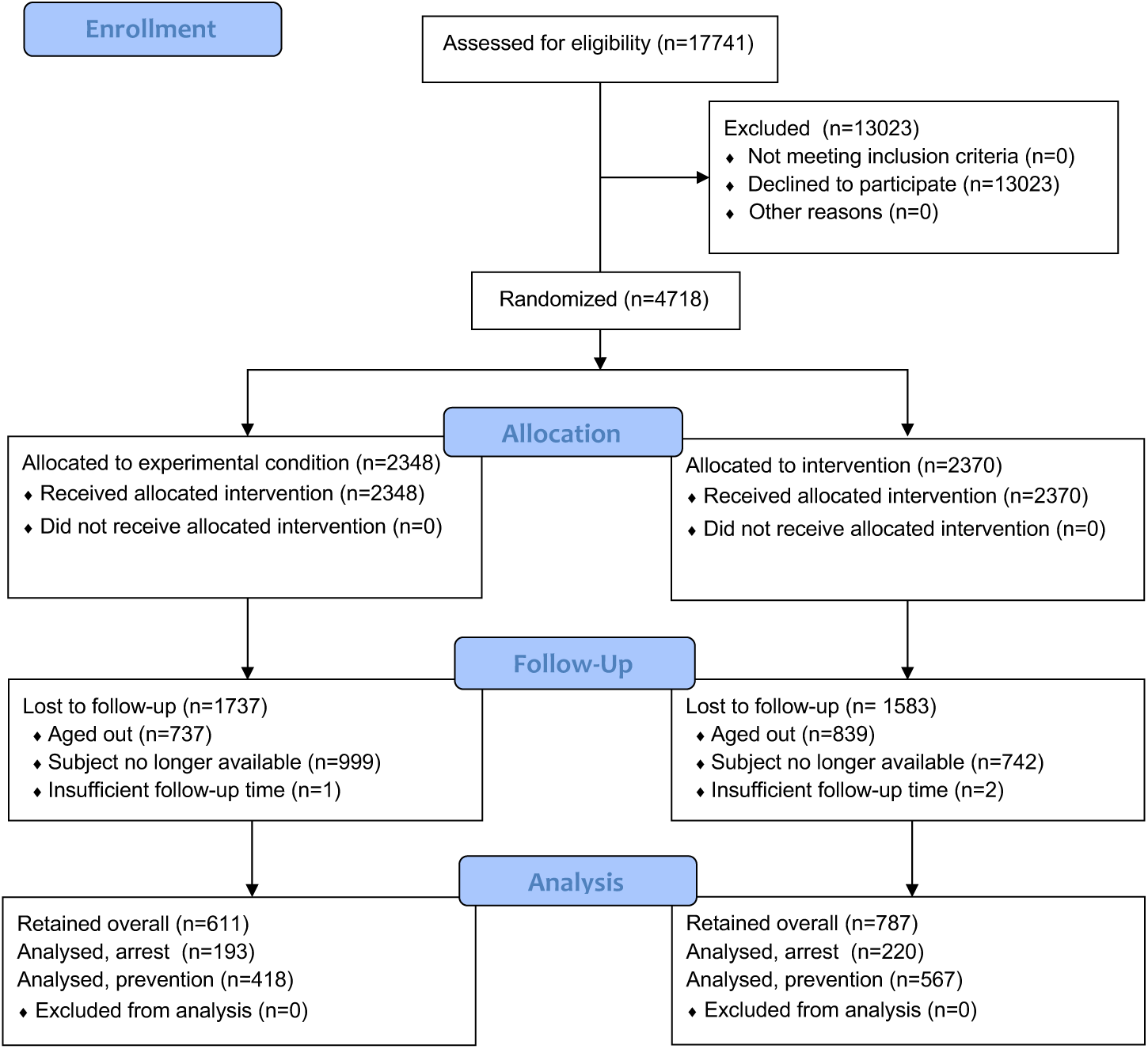
Study flow diagram

The baseline prevalence of sealants on any tooth prior to study intervention was 11% (N=156), with an untreated decay prevalence of 29.5% (Table 1). Hispanic/Latino and black children comprised 63% of the analytic sample and 54% were female. The average time that elapsed from baseline to follow-up for the analytic sample was 718 days. The intraclass correlation coefficients for caries arrest and prevention were 0.034 and 0.0031, respectfully.

**Table 1:** Baseline demographics overall and by treatment group (N=1398)

|  | **Overall** |  | **Simple** |  | **Complex** |  |
| --- | --- | --- | --- | --- | --- | --- |
|  | N/Mean | % / SD | N | % | N | % |
| Subjects | 1398 | 100 | 611 | 43.71 | 787 | 56.29 |
| Female | 753 | 53.86 | 321 | 52.54 | 432 | 54.89 |
| Race |  |  |  |  |  |  |
| Hispanic | 679 | 48.85 | 287 | 47.36 | 392 | 50 |
| Black | 208 | 14.96 | 98 | 16.17 | 110 | 14.03 |
| White | 29 | 2.09 | 17 | 2.81 | 12 | 1.53 |
| Asian | 24 | 1.73 | 14 | 2.31 | 10 | 1.28 |
| Multiple | 20 | 1.44 | 8 | 1.32 | 12 | 1.53 |
| Other | 11 | 0.79 | 7 | 1.16 | 4 | 0.51 |
| DK/Missing | 419 | 30.14 | 175 | 28.88 | 244 | 31.12 |
| Untreated decay | 413 | 29.54 | 193 | 31.59 | 220 | 27.95 |
| Sealants at baseline | 156 | 11.16 | 60 | 9.82 | 96 | 12.2 |
| Age at baseline | 6.63 | 1.21 | 6.57 | 1.23 | 6.68 | 1.19 |
| Decayed teeth | 0.69 | 1.39 | 0.72 | 1.36 | 0.66 | 1.42 |

The proportion of subjects who remained arrested for simple and complex groups (Table 2) were 0.56 (SE=0.04) and 0.46 (SE=0.42), respectively, for a difference of -0.11 (95% CI = -0.22, 0.01). Prevention rates amongst those without caries at baseline was 0.81 (SE=0.20) for the simple treatment and 0.82 (SE=0.17) for complex. The 95% confidence interval for the difference was (−0.04, 0.06). Simple group rates were non-inferior to those of complex. Non-inferiority for clinical outcomes is summarized in Figure 2.

**Table 2:** Non-inferiority results for arrest and prevention rates after two years

|  | **Simple** |  |  | **Complex** |  |  | **Difference** |  | **95% CI** |  |
| --- | --- | --- | --- | --- | --- | --- | --- | --- | --- | --- |
|  | N | mean | SE | N | mean | SE | mean | SE | Lower | Upper |
| Arrest | 193 | 0.56 | 0.04 | 220 | 0.46 | 0.42 | -0.11 | 0.06 | -0.22 | 0.01 |
| Prevention | 418 | 0.81 | 0.2 | 567 | 0.82 | 0.17 | 0.01 | 0.27 | -0.04 | 0.06 |

**Figure 2:**
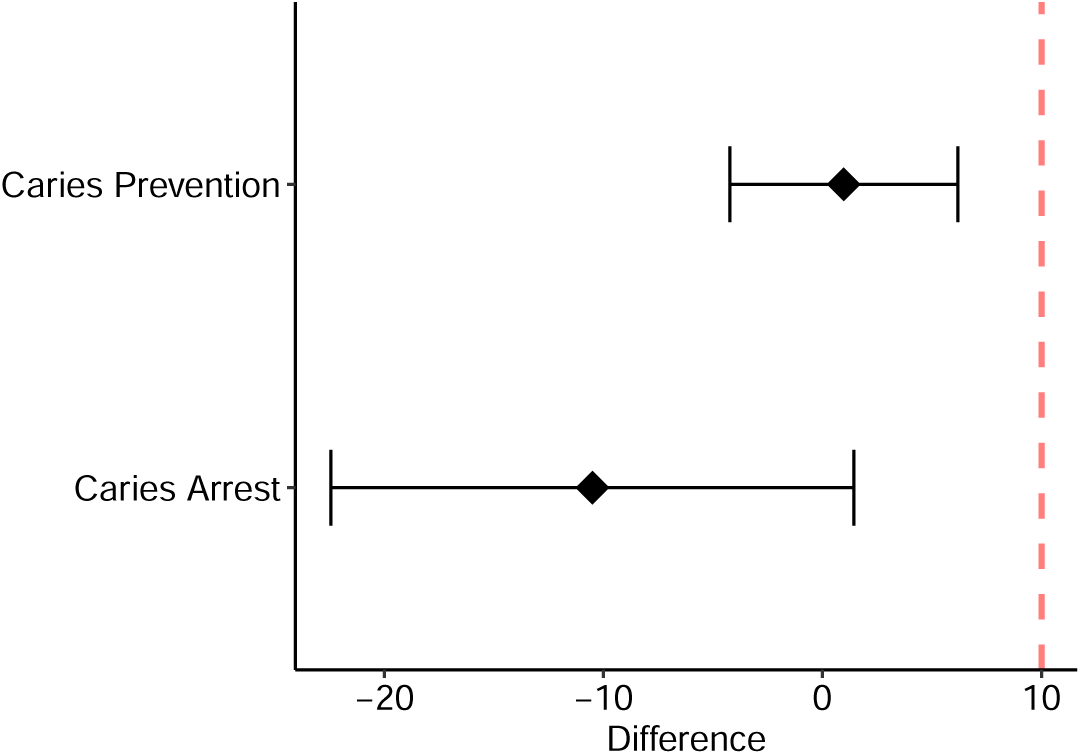
Non-inferiority plot for caries arrest and prevention at two years. The dashed red line denotes the non-inferiority limit.

## 4 Discussion

Without proper and timely prevention, dental caries and other oral diseases can lead to severe systemic infections [18], negatively impact oral health-related quality of life [19], and is associated with decreased student academic performance and school attendance [20]. To address the high rate of untreated caries in high-risk populations, the Centers for Disease Control and Prevention recommends school-based sealant programs, which have demonstrated clinical and cost effectiveness [21, 22, 23]. The Caried*Away* study explored the effectiveness of alternative, simple interventions that were applied using a school-based healthcare model.

In this pragmatic cluster randomized controlled trial, children who received a simple experimental treatment of fluoride varnish and silver diamine fluoride were non-inferior in the average two-year arrest and prevention of caries when compared to children receiving a standard comparator, a “complex” package of fluoride varnish, glass ionomer sealants, and atraumatic restorations. Our results support the utilization of silver diamine fluoride as an arresting and preventive agent for school-based oral health programs, complementing previous findings on non-restorative treatments in schools [24]. Both SDF and sealants demonstrated an approximate 80% prevention rate and 50% arrest rate after two years. These findings are comparable to those from more controlled, clinical studies, indicating no differences in the 6 and 12-month arrest rates comparing SDF versus ART [25]. Additionally, a review on the effect of SDF in preventing caries in the primary dentition showed significant reductions in the development of new caries versus placebo after 24 months, and was not more or less effective after twelve months compared to glass ionomer sealants [26].

The approximate two-year gap between initial treatment and follow-up coincided with municipal policies stemming from COVID-19 infection rates in New York City, with baseline observations being conducted over a six-month period from September 2019 to March 2020. On 16 March 2020, schools were closed citywide and dental offices suspended care except for emergency procedures. Schools remained closed to all school-based health programs throughout the 2020-2021 academic year. These restrictions meant that the original biannual data collection schedule for Caried*Away* could not be followed. Study activities resumed along with in-person learning beginning September 2021, however a substantial proportion of baseline subjects had aged out of the program and were not eligible for follow-up, resulting in our reported follow-up rate of 30%. Preliminary power calculations for Caried*Away* estimated a necessary sample size of 396 that was artificially increased by an a priori assumption of an intraclass correlation coefficient of 0.10, reflecting a moderate expectation of cluster correlation [15]. As we have shown, the actual degree of cluster correlation within schools is negligible.

New York City dental offices were authorized to reopen in June 2020 following the adoption of interim infection control and prevention guidelines, recommending the reduction of aerosol-generating procedures, the use of handpieces, air or water syringes, and ultrasonic scalers. However, school-based dental programs using these aersol-free methods were similarly prohibited from providing care. Due to these restrictions on preventive care, combined with the Caried*Away* subject population being specifically chosen because of their traditional lack of access to or utilization of routine dental care, it is unlikely that confounding dental treatments were received in the time between observations [27]. We further attempted to adjust for this in the analysis of arrest and prevention by simultaneously considering both untreated decay and any new fillings that were not present at baseline, which would be indicative of new disease incidence prior to follow-up.

The profound benefits of the caries arrest and prevention methods used in Caried*Away* offer substantial opportunities for optimizing oral health care and oral health in general, including sequential treatment protocols (e.g., silver diamine fluoride as an initial treatment with subsequent application of glass ionomer) and individual treatments for mixed dentition to reduce care delivery time (e.g., silver diamine on deciduous teeth and glass ionomer on permanent teeth). More broadly, these results counter current accepted guidance for school-based caries prevention, which recommends the use of sealants for first and second molars for children aged 6-12 years. Integration of newly recognized essential oral medicines can potentially dramatically increase the reach and effectiveness of school-based caries prevention and opens a new frontier for oral health providers.

## Data Availability

The datasets generated and/or analysed during the current study are not publicly available due to the active nature of the trial but are available from the corresponding author on reasonable request.

## 5 Additional information

### 5.2 Author contributions

RRR and RN conceived of the study. TBG provided treatments, collected clinical data, and co-directed clinical team activities. RRR performed all statistical analyses and wrote the manuscript. All authors critically reviewed the manuscript and provided edits. All authors read and approved the final manuscript.

